# Shifting From an Expected to an Opportunistic Pathogen: Characteristics and Trends of Infant Late and Very Late Onset Group B streptococcal (GBS) infection in a Canadian city over a 27-year period

**DOI:** 10.1101/2025.11.03.25339435

**Authors:** Isoken Isah, Sneha Suresh, Gregory J. Tyrrell, Manoj Kumar, Joan L. Robinson

## Abstract

**Background:** Despite decades of study, the characteristics of infants with invasive group B streptococcus (GBS) late onset disease (LOD) (onset day 7 to 89 of life) and in particular very late onset disease (VLOD) (after day 89 of life) are not well described.

**Materials and Methods:** This was a retrospective cohort study of infants hospitalized in four Edmonton hospitals April 1, 1994 through June 30, 2022 with LOD or VLOD GBS invasive disease. Data were collected on demographics, date of onset of GBS infection, clinical manifestations and outcomes.

**Results:** There were 115 episodes of LOD in 111 infants of which 49 (45%) were preterm. Onset was on median day 27 (IQR 19-40.5) of life. All but one infant was bacteremic while 38 (34%) had proven and 17 (15%) had possible GBS meningitis. Five (5%) died before hospital discharge with all deaths presumed to be due to GBS. There were 11 episodes of VLOD in 11 infants (7 [64%] preterm) presenting on median day 116 (IQR 103-138) (range 93-207) of life. Three (27%) had GBS meningitis. All survived to discharge. Infants with VLOD were less likely to be born vaginally and more likely to be mechanically ventilated during their birth hospitalization and have invasive disease with other pathogens than were infants with LOD. Serotype III accounted for 78% of LOD and 64% of VLOD cases.

**Conclusion:** GBS remains a significant cause of infant morbidity and mortality. There is increasing evidence that GBS may sometimes be an opportunistic pathogen in infants over 90 days of age.

## Introduction

*Streptococcal agalactiae,* or group B streptococcus (GBS) is part of rectovaginal flora for approximately 10 to 35% of women (1). Carriage in pregnant persons can lead to neonatal invasive GBS disease; this was a rare entity until the 1970’s when GBS became the leading cause of neonatal sepsis. The reasons for this epidemiologic shift are not clear.

GBS early onset disease (EOD) is transmitted from rectovaginal colonization during labor or at delivery and presents in the first 6 days of life. Risk factors for EOD include maternal GBS bacteriuria (an indicator of heavy colonization), chorioamnionitis (as the uterus may contain a high concentration of GBS), prolonged rupture of membranes, preterm delivery (which can be due to symptomatic or subclinical GBS chorioamnionitis) and birth of a previous infant with GBS infection (presumably because the mother does not produce effective GBS antibodies) (1). Intrapartum antibiotic prophylaxis (IAP) markedly decreases the incidence of EOD (1).

GBS late-onset disease (LOD) occurs after day 6 and very late onset disease (VLOD) after day 89 of life. The pathogenesis of GBS acquisition and infection in LOD and VLOD are not established, with some studies implicating breast milk as a source of LOD (2,6). IAP does not change the incidence of LOD (1) so has not been studied with VLOD.

The objective of this study was to describe and compare the incidence and characteristics of LOD and VLOD in a cohort of neonates in a Canadian city over a prolonged period.

## Materials and Methods

This was a retrospective study of the cohort of infants born in Edmonton or St. Albert and hospitalized in one of the four Edmonton hospitals April 1, 1994 through June 30, 2022 that had GBS isolated from a sterile site (excluding urine) after day 6 of life. LOD was defined as presentation on day 7 to 89 while VLOD as presentation on day 90 to 364 of life. The GBS prenatal protocol during these years included rectovaginal screening at 35 to 37 weeks gestation with intrapartum antibiotics for all who screened positive and for unscreened persons with risk factors for GBS transmission.

Cases were selected via health records search of inpatients discharges up to 12 months of age with discharge ICD9CM/ICD10CA diagnostic codes that would capture GBS sepsis (S1 Table). Eligibility was confirmed by chart review. Cross referencing of LOD cases was done through a provincial microbiological database of all infants with GBS isolated from a sterile site up to day 89 of life. Cross referencing was not possible for VLOD cases.

Charts were reviewed and data collected on demographics, date of onset of GBS infection, clinical manifestations and outcomes (S2 Table).

Cerebrospinal fluid (CSF) pleocytosis was defined as > 20 × 10^9^/L WBCs using a ratio of 1000RBCs:1WBC.

Ethics approval was obtained from the University of Alberta Health Research Ethics Board (Pro00059616) who waived the need for parental consent. They approved entry of anonymized data and date of birth into REDCap.

### Data analysis

The data for the characteristics of LOD and VLOD cases was compared to identify any differentiating features among these groups. Chi-square tests were used to compare dichotomous variables, with the use of Fisher’s exact test to estimate statistical significance if the expected cell frequency were <5 for any of the cells in 2×2 tables. Data were analyzed using OpenEpi, an open-source software (Available at www.openepi.com). Data on continuous variables are provided as means with standard deviation (SD) or medians with inter-quartile range, depending on the characteristics of the data distribution. Incidence of LOD and VLOD per 1000 live births was calculated with live birth denominator data from Statisa 2025 (https://www.statista.com/statistics/578589/number-of-births-in-alberta-canada/). Figures were created with the assistance of ChatGPT (OpenAI, San Francisco, CA).

### Capsular polysaccharide (CPS) typing

Neonatal invasive GBS disease is a notifiable disease in Alberta, requiring GBS isolates to be submitted to the Alberta Public Health Laboratory for CPS typing (2). Prior to January 2017, CPS typing was performed using a double immunodiffusion method with CPS-specific antisera (3). From 2017 on, a real-time PCR assay was used (4).

## Results

Screening of discharge codes yielded 842 infants of which 122 had confirmed LOS.

### Late onset disease (day 7 to 89 of life)

There were 111 infants with 115 episodes of culture-confirmed GBS LOD (37 with positive blood and CSF cultures, 1 with positive CSF cultures only and 77 with positive blood cultures). Onset was on median day 27 (IQR 19-40.5) of life; 47 (42%) were male and 49 (45%) were preterm (data on gestation missing for 2 cases) (Table 1). At the time of GBS infection, 69 (62%) were receiving breast milk (66 mother’s own milk and 3 donor human milk) while 5 (5%) required parenteral nutrition.

**Table 1.**
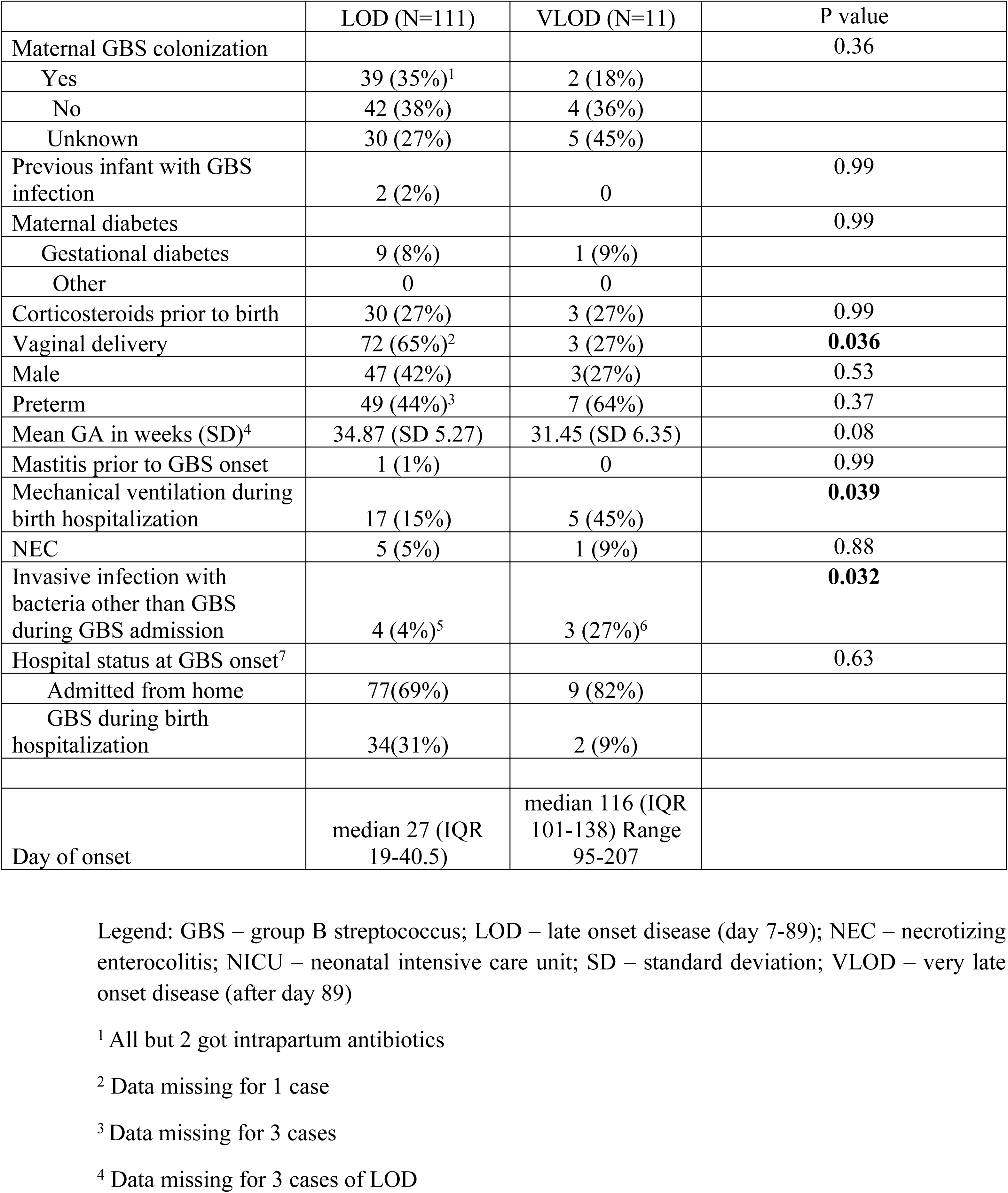
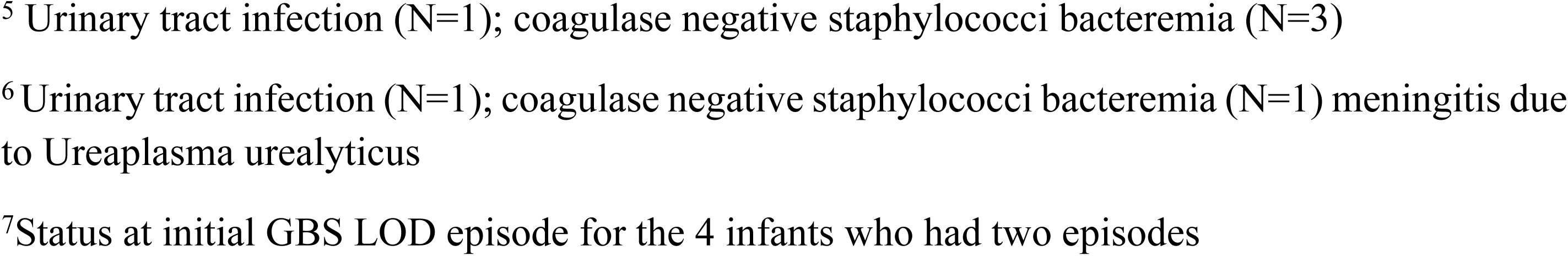
– Comparison of LOD and VLOD cases.

|  | LOD (N=111) | VLOD (N=11) | P value |
| --- | --- | --- | --- |
| Maternal GBS colonization |  |  | 0.36 |
| Yes | 39 (35%)<sup>1</sup> | 2 (18%) |  |
| No | 42 (38%) | 4 (36%) |  |
| Unknown | 30 (27%) | 5 (45%) |  |
| Previous infant with GBS infection | 2 (2%) | 0 | 0.99 |
| Maternal diabetes |  |  | 0.99 |
| Gestational diabetes | 9 (8%) | 1 (9%) |  |
| Other | 0 | 0 |  |
| Corticosteroids prior to birth | 30 (27%) | 3 (27%) | 0.99 |
| Vaginal delivery | 72 (65%)<sup>2</sup> | 3 (27%) | **0.036** |
| Male | 47 (42%) | 3 (27%) | 0.53 |
| Preterm | 49 (44%)<sup>3</sup> | 7 (64%) | 0.37 |
| Mean GA in weeks (SD)<sup>4</sup> | 34.87 (SD 5.27) | 31.45 (SD 6.35) | 0.08 |
| Mastitis prior to GBS onset | 1 (1%) | 0 | 0.99 |
| Mechanical ventilation during birth hospitalization | 17 (15%) | 5 (45%) | **0.039** |
| NEC | 5 (5%) | 1 (9%) | 0.88 |
| Invasive infection with bacteria other than GBS during GBS admission | 4 (4%)<sup>5</sup> | 3 (27%)<sup>6</sup> | **0.032** |
| Hospital status at GBS onset<sup>7</sup> |  |  | 0.63 |
| Admitted from home | 77(69%) | 9 (82%) |  |
| GBS during birth hospitalization | 34(31%) | 2 (9%) |  |
| Day of onset | median 27 (IQR 19-40.5) | median 116 (IQR 101-138) Range 95-207 |  |

Legend: GBS – group B streptococcus; LOD – late onset disease (day 7-89); NEC – necrotizing
enterocolitis; NICU – neonatal intensive care unit; SD – standard deviation; VLOD – very late
onset disease (after day 89)
<sup>1</sup> All but 2 got intrapartum antibiotics
<sup>2</sup> Data missing for 1 case
<sup>3</sup> Data missing for 3 cases
<sup>4</sup> Data missing for 3 cases of LOD
<sup>5</sup> Urinary tract infection (N=1); coagulase negative staphylococci bacteremia (N=3)
<sup>6</sup> Urinary tract infection (N=1); coagulase negative staphylococci bacteremia (N=1) meningitis due to *Ureaplasma urealyticus*

At least one CSF was obtained from 108 of the 115 episodes (94%); 38 (35%) were culture-positive (19 obtained prior to antibiotics, 19 after at least one dose antibiotics) and 70 (65%) were culture-negative (22 obtained prior to antibiotics, 42 after at least one dose antibiotics and 6 with unknown timing). For 17 of the CSFs obtained after antibiotics or with unknown timing, there was CSF pleocytosis so the total number of episodes with confirmed (N=38) or possible (N=17) meningitis was 55 (48%). Eight cases had more than one positive CSF culture with these occurring day 1 (N=1), day 2 (N=2), day 3 (N=2) day 4 (N=2) and day 13 (N=1) after the initial positive culture.

In addition to bacteremia and/ or meningitis, local sites of infection included adenitis (N=2), omphalitis (N=1), cellulitis (N=3), adenitis and cellulitis (N=1), and omphalitis and cellulitis (N=1). Another 7 infants had positive urine cultures for GBS in addition to positive blood or CSF cultures. There were no musculoskeletal infections.

Three of the 111 infants had EOD before the episode of LOD including one infant who went on to have two episodes of LOD. That infant was born at 36 weeks gestation and had GBS bacteremia without a focus on days 6, 19 and 45 of life. No explanation was established but breastfeeding was stopped after the second recurrence as breast milk and infant throat culture grew GBS. Another three infants had two episodes of LOD. A 28-week GA infant had meningitis on day 13 and bacteremia with no focus on day 43 of life (5 days after a 25-day course of antibiotics ended). A 29-week GA infant had bacteremia with no focus on day 39 and day 56 of life (7 days after a 10-day course of antibiotics ended). The third infant born at 27 weeks GA had LOD days 19 and day 36 of life (4 days after a 13-day course of antibiotics for bacteremia ended) with possible meningitis in the second episode (CSF pleocytosis in a culture-negative bloody tap obtained after antibiotics were given).

Fourteen of the 111 infants (13%) were discharged on anticonvulsants; 11 of these had confirmed and 2 had possible meningitis. Four infants had documented hearing loss, of whom 2 had meningitis. Three infants required ventriculoperitoneal shunts.

Five infants (5%) died before hospital discharge; 4 died on day 0 to 5 of treatment for GBS and one had withdrawal of care after treatment was completed.

The number of cases over time is shown in Figure 1.

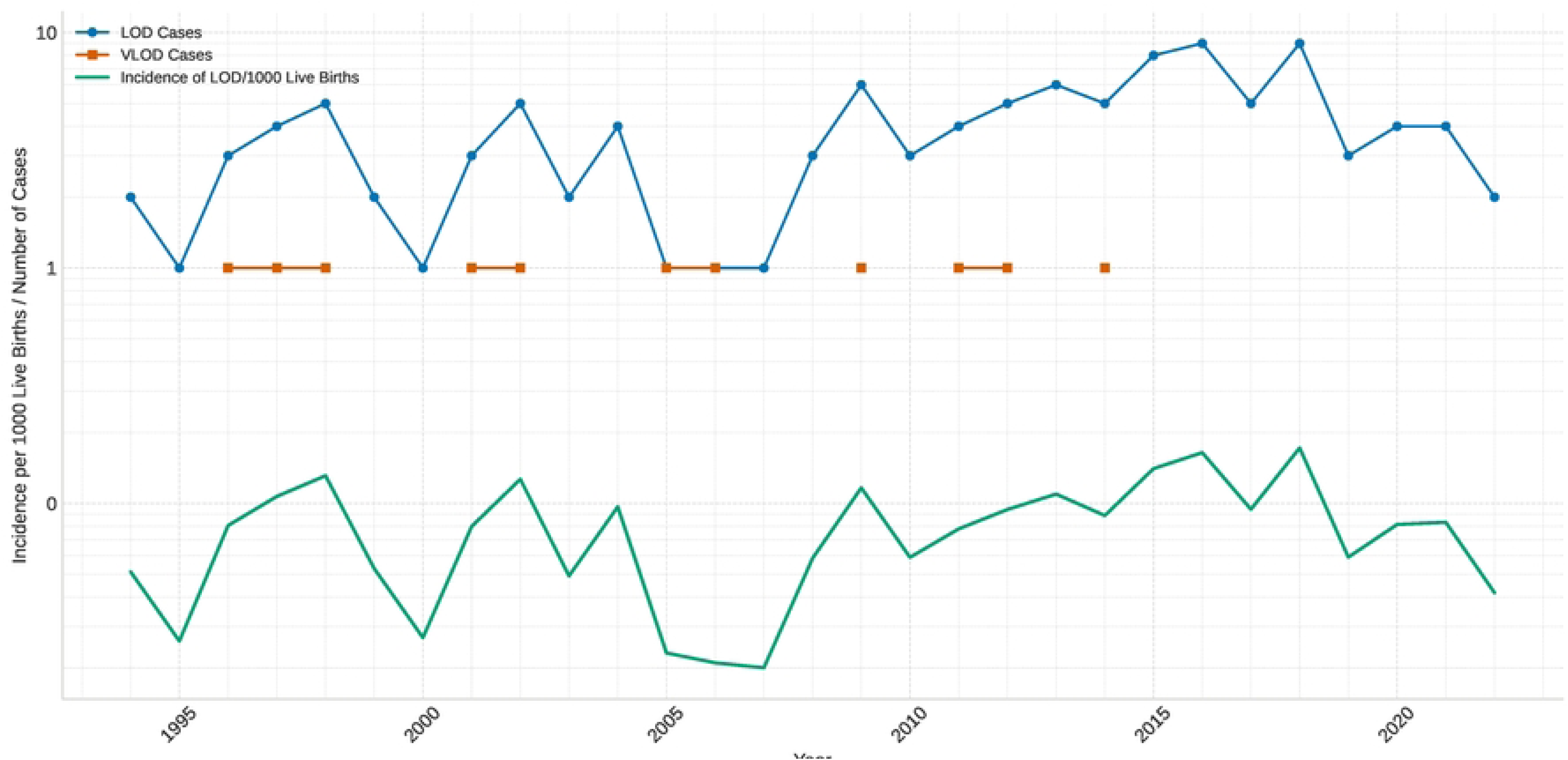
Figure 1.

Serotypes from 101 of the 111 infants are shown in Figure 2 with 78 (77%) being serotype III (For the four infants with two episodes of LOD, serotypes were identified only on the initial isolate). Data were omitted from one infant as serotype III was identified in blood and serotype V in CSF; it seems likely that one of these results was incorrect, but specimens are no longer available for retesting.

**Figure 2.**
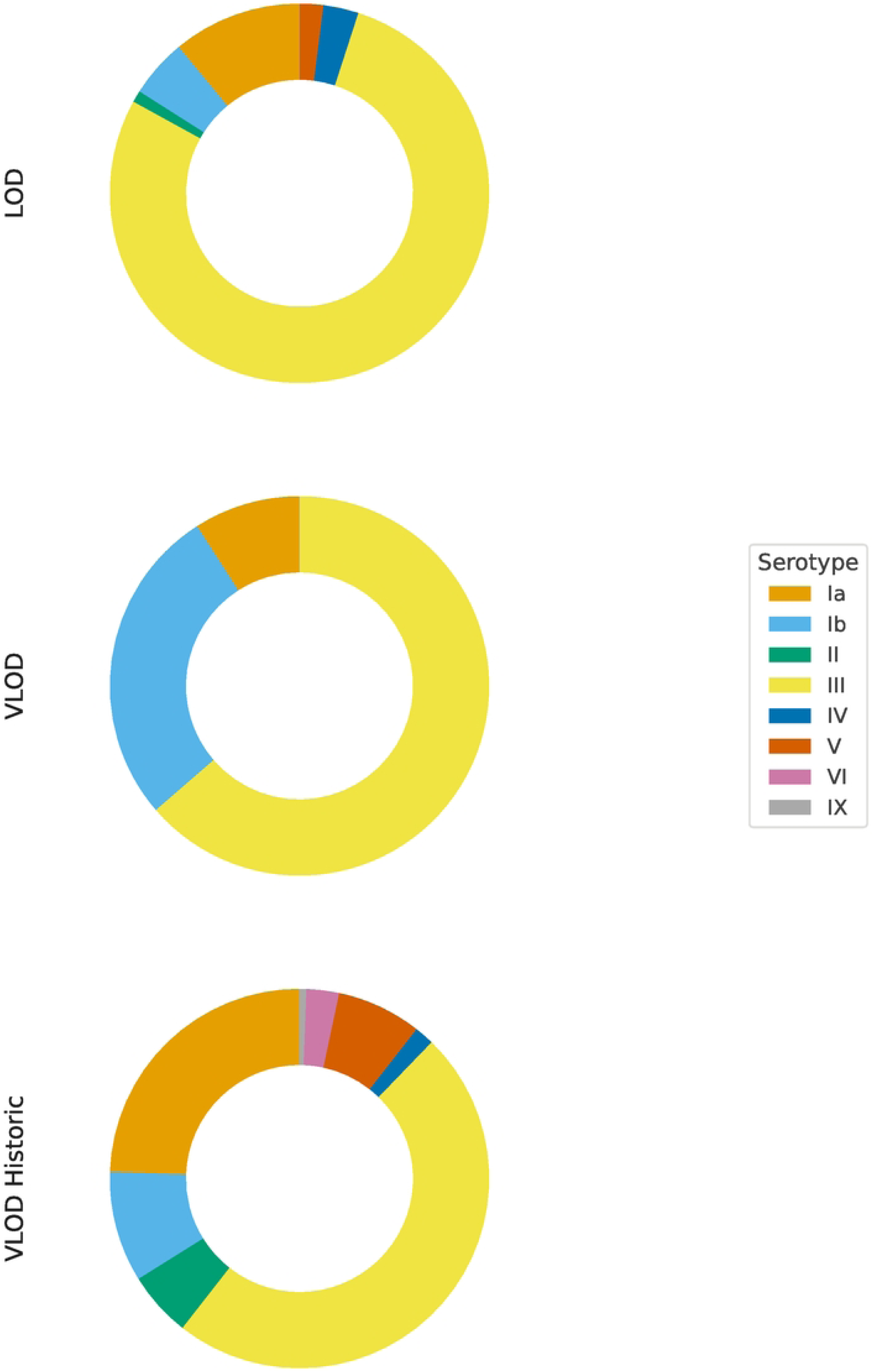
– GBS serotypes from LOD and VLOD cases in the current study and historic VLOD cases reported in the literature.

### Very late onset disease (day 90-364)

There were 11 VLOD cases presenting on median day 116 (IQR 103-138) (range 93-207) of life (Table 1). Three infants were male (27%) and 7 (64%) were preterm. None had been diagnosed with chronic medical conditions at the time of GBS onset.

CSF was obtained from all 11 cases. Three (27%) were culture-positive (2 obtained prior to antibiotics and 1 after at least one dose antibiotics) and 8 were culture-negative (3 obtained prior to antibiotics, 5 after at least one dose antibiotics). None of the 5 with negative cultures obtained after antibiotics had CSF pleocytosis.

Local sites of infection included 3 cases with cellulitis, none of whom had meningitis.

One infant (with meningitis) was discharged on anticonvulsants. Six had normal hearing when tested and 5 were not tested. None had prior or subsequent GBS infections. All survived to discharge.

Serotypes were Ia (N=1), Ib (N=3), and III (N=7).

The number of cases over time is shown in Figure 1 with there never being more than one VLOD case per year.

### Comparison of LOD to VLOD cases

LOD cases were more likely to be born vaginally, less likely to have required mechanical ventilation during their birth hospitalization, and less likely to have invasive infections other than GBS when compared to VLOD cases (Table 1). The proportion of isolates that were serotype III did not differ between LOD and VLOD cases (P=0.51).

## Discussion

Over a 27-year period in Edmonton hospitals, there was a mean of 4.3 infants per year with GBS invasive disease with onset after day 6 of life (range 1 to 10 infants). Approximately 10% had VLOD (11 of 122). Statistically significant differences between LOD and VLOD were a higher incidence of vaginal delivery with LOD, and a higher incidence of mechanical ventilation during birth hospitalization and invasive infection with bacteria other than GBS noted with VLOD.

For LOD, it is not clear when transmission most commonly occurs, i.e., intrapartum, from the mother after delivery (from GBS on her hands or from breastmilk), from nosocomial transmission, or from the community. In one novel case in the literature, identical twins (36 weeks GA) who were initially breastfed (mode of feeding at the time of presentation was not clarified) presented on the same day with GBS meningitis at 14 weeks of age (5); clearly they had a common high-grade exposure which was never identified. The incidence of LOD in Alberta increased from 0.15 to 0.41 per 1000 live births from 2003 to 2020 for unclear reasons (2, 6).

A 2022 systematic review of LOD reported an odds ratio (OR) of 2.67 (95% confidence interval (CI): 2.07-3.45) for maternal GBS colonization (7). Furthermore, a prospective cohort of 100 infants with LOD reported that 30 of 47 mothers (64%) had GBS colonization (8). In one study, IAP was associated with later presentation of LOD and milder disease (8). This all suggests that intrapartum transmission accounts for some LOD. However, in the systematic review, maternal fever (a marker of possible chorioamnionitis) and prolonged rupture of membranes were not risk factors for LOD (7) as one might expect them to be if the bulk of cases stemmed from intrapartum transmission.

One study showed that about 25% of infants are colonized with the maternal GBS strain by 8 weeks of age despite IAP (9). It is not clear whether low-level transmission still occurred at birth or whether GBS is commonly acquired from the mother after delivery. The source of post-partum acquisition might sometimes be breastmilk. GBS mastitis has been documented in only a small percentage of LOD cases (8) but it is possible that transmission occurs in the absence of mastitis. A study from Australia of 92 LOD cases and 368 controls found no link between breastfeeding and LOD (10), suggesting that breast milk is not a common source of LOD. Neonatal colonization may lead to colonization of the breast and positive breastmilk cultures (1) and it remains possible that breast milk is never or rarely a source of LOD.

Nosocomial LOD occurs, mainly in neonatal intensive care units (11) with it not being clear how often the source is another infant versus colonized staff. It is difficult to prove the mechanism or incidence of community transmission leading to GBS infection.

Even less is known about the source of GBS in VLOD than in LOD. Our review of the literature for GBS case series identified 352 cases in 13 studies dating back to 1995 (Table 2). Three were population based studies that reported the incidence per 1000 live births: i) LOD 0.36 (95% CI: 0.31–0.42) and VLOD 0.03 (95% CI: 0.01–0.04) in New Zealand 2012-2021(12), ii) LOD 0.20 (95% CI: 0.17–0.23) and VLOD 0.012 (95% CI: 0.007–0.021) in Norway 1996-2012 (13), and iii) LOD 0.12 (95% CI 0.11–0.14), and VLOD 0.01 (95% CI 0.01–0.02) in Japan 2011-2015 (14). The upper age limit for VLOD was 6 months in the New Zealand study and 12 months in the other two studies. Combining data from the 8 studies that included cases up to 1 year of age, there were 2242 cases of EOD, 2668 of LOD and 265 of VLOD – again indicating that as in our study, VLOD cases account for about 10% of GBS cases after 6 days of age. For the 10 studies that reported GA, 82 of 208 infants (39%) were preterm. Six studies reported clinical presentation with 61 of 175 infants (35%) having meningitis, 94 (54%) having bacteremia with no focus mentioned in the manuscript and 20 (11%) having bacteremia with a focus: urinary tract infection (N=7); cellulitis (n=4); septic arthritis (N=3); central line associated blood stream infection (N=2); infective endocarditis (N=2) and a bullous rash (N=1).

**Table 2.**
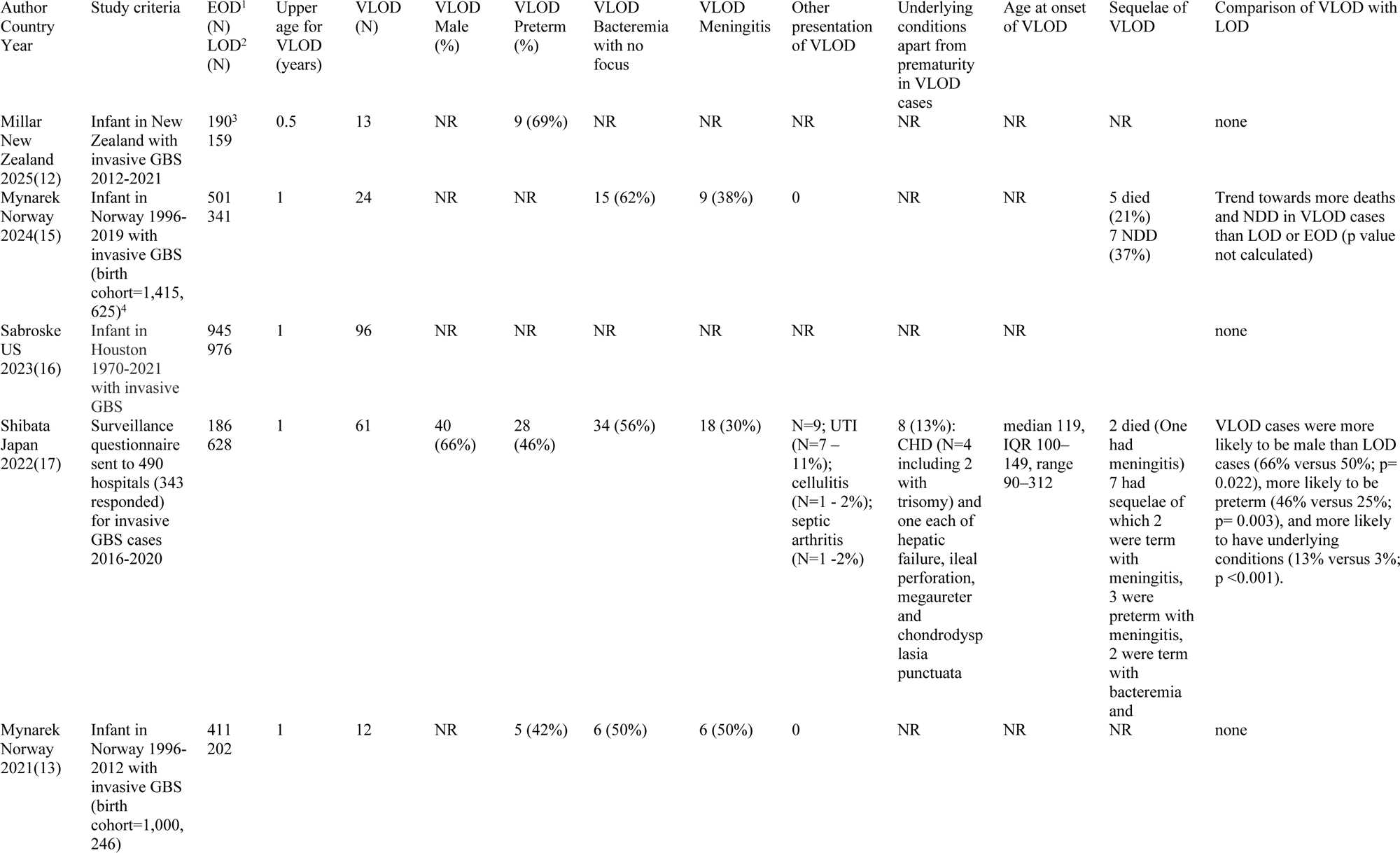
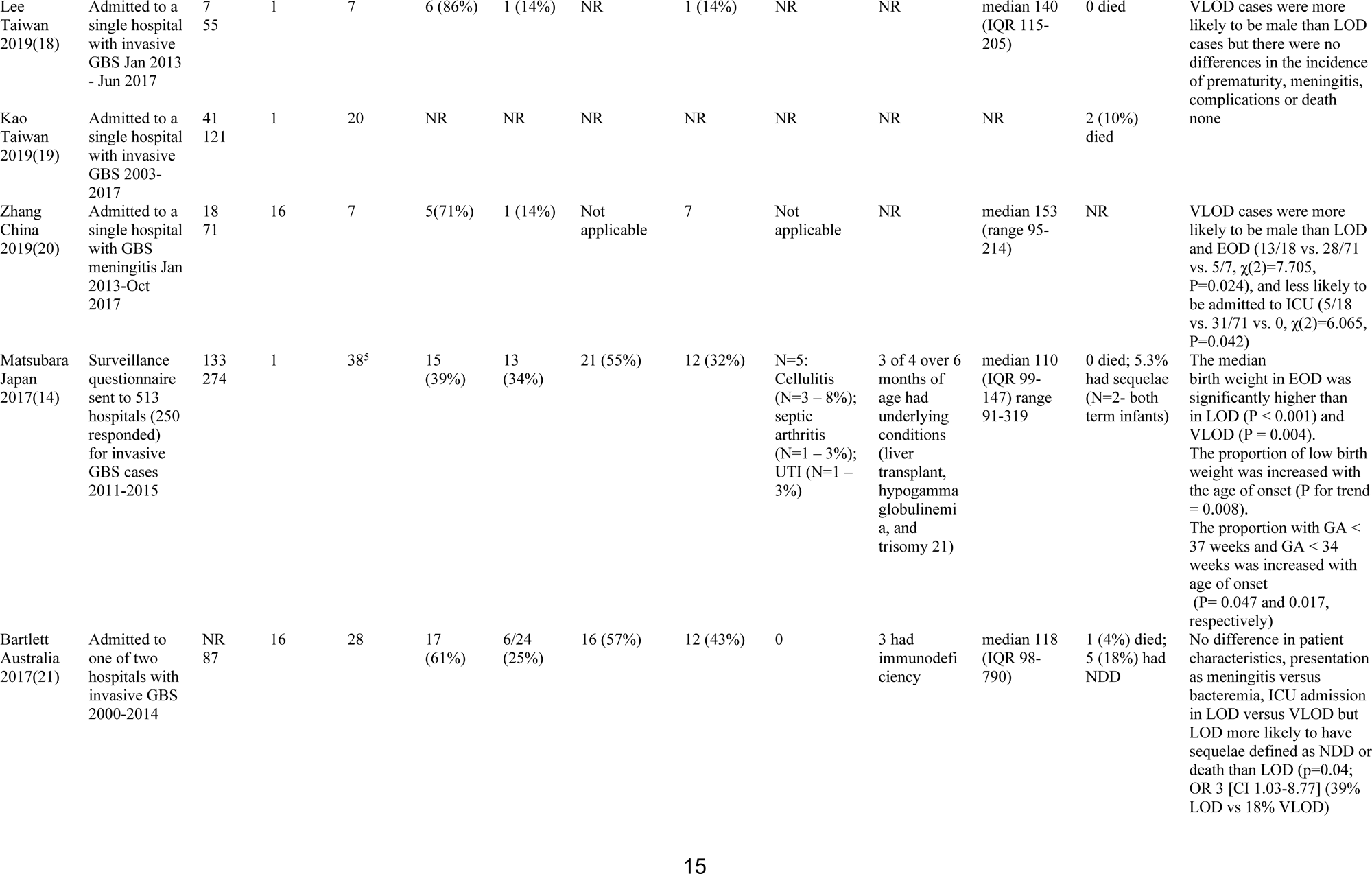
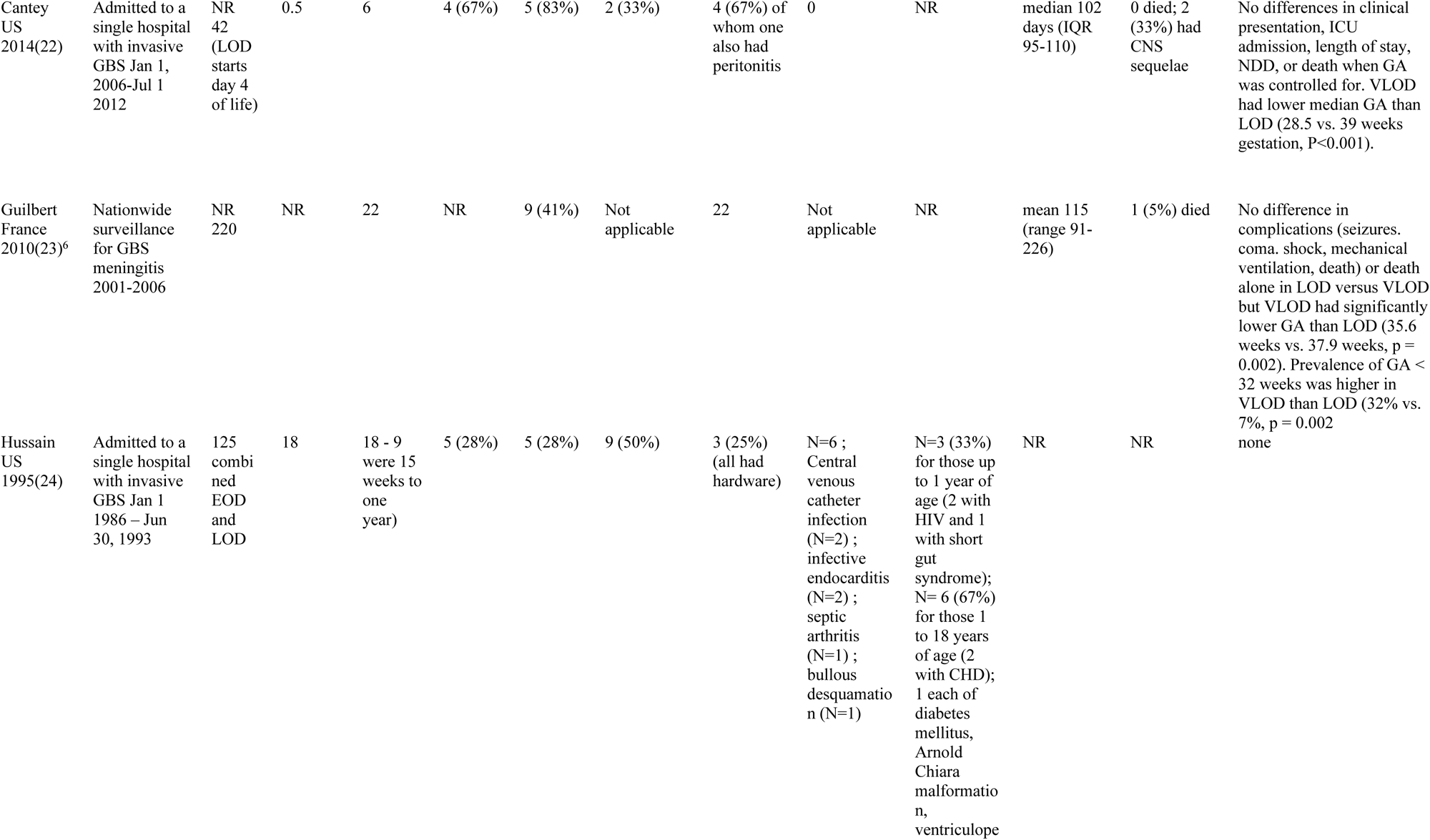
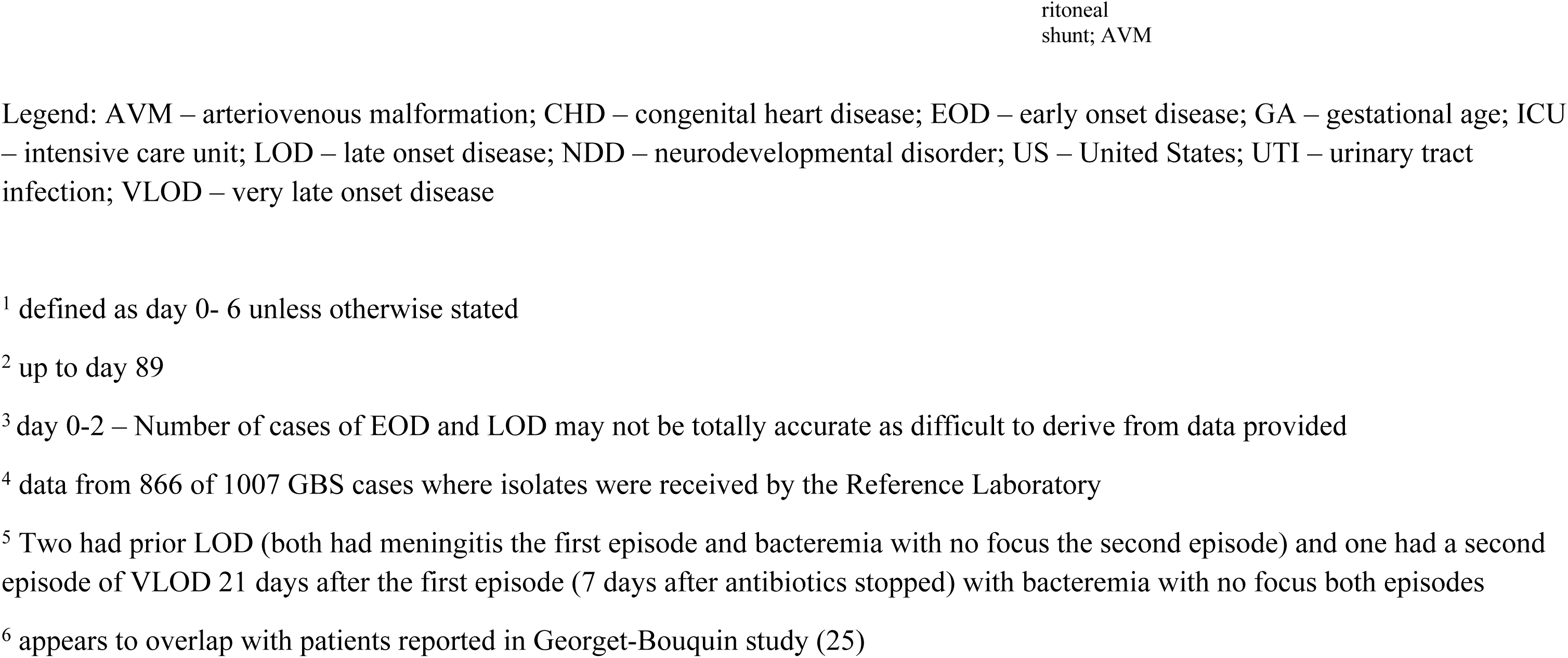
– Literature review of case series of VLOD due to GBS.

In the literature review, a risk factor for VLOD as compared to LOD +/-EOD included prematurity in 4 studies (14, 17, 22, 23) and male gender in 3 studies (17, 18, 20) (Table 2). However, combining data from the 6 studies that reported gender with our study, 85 of 169 VLOD cases were male (50%), so it seems unlikely that there is a true gender predominance. Another risk factor for VLOD in one study was the presence of underlying disorders (17). A major limitation is that underlying disorders would often not yet be diagnosed at the time of LOD or VLOD presentation so are not reliably mentioned in published studies. Immune deficiency is thought to lead to some cases of VLOD. In the review of the literature, at least 2 of the 352 VLOD cases had human immunodeficiency virus and one study reported that 3 of 28 cases had “immunodeficiency”. There is a case report of a child with IRAK-4 deficiency who presented with GBS meningitis at 5 months of age (26). A recurrent case of VLOD has also been reported in a child with thoracic congenital lymphatic dysplasia (which is associated with CD4 lymphopenia and hypogammaglobulinemia) with two episodes of bacteremia 5 months apart starting at 2 years of age (27). In our cohort, VLOD infants were more likely to have invasive infections other than GBS, which could signal a defect in host defense. However immune deficiency was not limited to VLOD cases, as one of our infants with LOD (GBS meningitis at 15 days of age) went on to be diagnosed with an Nf kappa B pathway immune deficiency that also affected toll-like receptor (TLR) function similar to IRAK-4 deficiency. The correlation between inborn errors of immunity that affect TLR function and VLOD/LOD GBS disease needs further study.

In the literature review, in terms of severity of illness, one study reported at least a trend towards a higher risk of neurodevelopmental disorders (NDD) and of death with VLOD than with EOD or LOD (P value not provided) (15) while another study reported the opposite – a decreased risk of NDD or death with VLOD than with LOD (21) (Table 2). Another study reported a lower risk of ICU admission with VLOD than with LOD and EOD combined (20).

It has been postulated that VLOD has the same origins and clinical course as LOD, occurring in infants who were typically more preterm so present at an older chronological age (22). However, this theory does not account for cases in term babies, which accounted for 61% of cases in the literature review (Table 2) and about one-third of cases in the current study.

It is possible that the pathogenicity of certain serotypes of GBS predisposes to later onset disease. Serotype III is the most common cause of both EOD and LOD infant invasive GBS (26). Sequence type 17 and its associated virulence factor HvGA are associated with infant invasive GBS, with the majority of these cases being serotype III (26); unfortunately, such testing was not available for the isolates from the current study.

Serotyping was reported for 180 of the 352 VLOD cases in the literature review with 87 (48%) being serotype III, while in our cohort 64% of VLOD cases were serotype III (Figure 2). In the literature review, some VLOD cases were with t uncommon serotypes (VI: N=5; XI: N=1) and there is a case report of VLOD meningitis due to serotype IX presenting at 120 days of age (28). This may further support the hypothesis that a defect in host defense is sometimes the culprit in VLOD, as less pathogenic serotypes of GBS are responsible for VLOD cases.

Our study shows that VLOD accounts for about 10% of GBS disease after 6 days of age. While prematurity is a risk factor in previous studies, further exploration into defects in infant microbial defense, particularly innate immune defects, is warranted. It seems possible that infant GBS shifts from being an expected pathogen to an opportunistic one with maturity and that an immune disorder should be considered in infants with VLOD.

## Author contributions

Isoken Isah – Data Curation; Writing – Review & Editing

Sneha Suresh – Conceptualization; Methodology; Supervision; Visualization, Writing – Review & Editing

Gregory Tyrrell-Methodology; Data Curation; Writing – Review & Editing

Manoj Kumar – Formal Analysis; Writing – Review & Editing

Joan L. Robinson – Methodology; Supervision; Writing – Original Draft Preparation

## Data Availability

Data cannot be shared publicly because of the confidentiality of patient data in a relatively small data set. Data are available from the University of Alberta Institutional Data Access / Ethics Committee (contact via 780-492-0302) for researchers who meet the criteria for access to confidential data.

---

**S1 Table.** ICD9CM/ICD10CA diagnostic codes.

**S2 Table.** Case report form.

## References

1. Coggins SA, Puopolo KM. Neonatal Group B Streptococcus Disease. Pediatr Rev. 2024;45(2):63–73.

2. Ma A, Thompson LA, Corsiatto T, Hurteau D, Tyrrell GJ. Epidemiological Characterization of Group B Streptococcus Infections in Alberta, Canada: An Update from 2014 to 2020. Microbiol Spectr. 2021;9(3):e0128321.

3. Lancefield RC, McCarty M, Everly WN. Multiple mouse-protective antibodies directed against group B streptococci. Special reference to antibodies effective against protein antigens. J Exp Med. 1975;142(1):165–79.

4. Alhhazmi A, Pandey A, Tyrrell GJ. Identification of Group B Streptococcus Capsule Type by Use of a Dual Phenotypic/Genotypic Assay. J Clin Microbiol. 2017;55(9):2637–50.

5. Sherman G, Lamb GS, Platt CD, Wessels MR, Chochua S, Nakamura MM. Simultaneous Late, Late-Onset Group B Streptococcal Meningitis in Identical Twins. Clin Pediatr (Phila). 2023;62(2):96–9.

6. Alhhazmi A, Hurteau D, Tyrrell GJ. Epidemiology of Invasive Group B Streptococcal Disease in Alberta, Canada, from 2003 to 2013. J Clin Microbiol. 2016;54(7):1774–81.

7. Karampatsas K, Davies H, Mynarek M, Andrews N, Heath PT, Le Doare K. Clinical Risk Factors Associated With Late-Onset Invasive Group B Streptococcal Disease: Systematic Review and Meta-Analyses. Clin Infect Dis. 2022;75(7):1255–64.

8. Berardi A, Rossi C, Lugli L, Creti R, Bacchi Reggiani ML, Lanari M, et al. Group B streptococcus late-onset disease: 2003-2010. Pediatrics. 2013;131(2):e361–8.

9. Berardi A, Rossi C, Creti R, China M, Gherardi G, Venturelli C, et al. Group B streptococcal colonization in 160 mother-baby pairs: a prospective cohort study. J Pediatr. 2013;163(4):1099–104.e1.

10. Ching NS, Buttery JP, Lai E, Steer AC, Standish J, Ziffer J, et al. Breastfeeding and Risk of Late-Onset Group B Streptococcal Disease. Pediatrics. 2021;148(3).

11. Collin SM, Lamb P, Jauneikaite E, Le Doare K, Creti R, Berardi A, et al. Hospital clusters of invasive Group B Streptococcal disease: A systematic review. J Infect. 2019;79(6):521–7.

12. Millar JR, Anglemyer A, Werno A, Austin NC, Walls T. Epidemiology of Infant Group B Streptococcus Infection in New Zealand: A 10-Year Retrospective Study. Pediatr Infect Dis J. 2025.

13. Mynarek M, Bjellmo S, Lydersen S, Afset JE, Andersen GL, Vik T. Incidence of invasive Group B Streptococcal infection and the risk of infant death and cerebral palsy: a Norwegian Cohort Study. Pediatr Res. 2021;89(6):1541–8.

14. Matsubara K, Hoshina K, Kondo M, Miyairi I, Yukitake Y, Ito Y, et al. Group B streptococcal disease in infants in the first year of life: a nationwide surveillance study in Japan, 2011-2015. Infection. 2017;45(4):449–58.

15. Mynarek M, Vik T, Andersen GL, Brigtsen AK, Hollung SJ, Larose TL, et al. Mortality and neurodevelopmental outcome after invasive group B streptococcal infection in infants. Dev Med Child Neurol. 2024;66(1):125–33.

16. Sabroske EM, Iglesias MAS, Rench M, Moore T, Harvey H, Edwards M, et al. Evolving antibiotic resistance in Group B Streptococci causing invasive infant disease: 1970-2021. Pediatr Res. 2023;93(7):2067–71.

17. Shibata M, Matsubara K, Matsunami K, Miyairi I, Kasai M, Kai M, et al. Epidemiology of group B streptococcal disease in infants younger than 1 year in Japan: a nationwide surveillance study 2016-2020. Eur J Clin Microbiol Infect Dis. 2022;41(4):559–71.

18. Lee CC, Hsu JF, Prasad Janapatla R, Chen CL, Zhou YL, Lien R, et al. Clinical and Microbiological Characteristics of Group B Streptococcus from Pregnant Women and Diseased Infants in Intrapartum Antibiotic Prophylaxis Era in Taiwan. Sci Rep. 2019;9(1):13525.

19. Kao Y, Tsai MH, Lai MY, Chu SM, Huang HR, Chiang MC, et al. Emerging serotype III sequence type 17 group B streptococcus invasive infection in infants: the clinical characteristics and impacts on outcomes. BMC Infect Dis. 2019;19(1):538.

20. Zhang XX, Geng ZX, Zhu L, Li MH, Wang YJ, Qian SY, et al. [Clinical analysis of children with group B streptococcal meningitis in 2013-2017 in a single center]. Zhonghua Er Ke Za Zhi. 2019;57(6):452–7.

21. Bartlett AW, Smith B, George CR, McMullan B, Kesson A, Lahra MM, et al. Epidemiology of Late and Very Late Onset Group B Streptococcal Disease: Fifteen-Year Experience From Two Australian Tertiary Pediatric Facilities. Pediatr Infect Dis J. 2017;36(1):20–4.

22. Cantey JB, Baldridge C, Jamison R, Shanley LA. Late and very late onset group B Streptococcus sepsis: one and the same? World J Pediatr. 2014;10(1):24–8.

23. Guilbert J, Levy C, Cohen R, Delacourt C, Renolleau S, Flamant C. Late and ultra late onset Streptococcus B meningitis: clinical and bacteriological data over 6 years in France. Acta Paediatr. 2010;99(1):47–51.

24. Hussain SM, Luedtke GS, Baker CJ, Schlievert PM, Leggiadro RJ. Invasive group B streptococcal disease in children beyond early infancy. Pediatr Infect Dis J. 1995;14(4):278–81.

25. Georget-Bouquinet E, Bingen E, Aujard Y, Levy C, Cohen R. [Group B streptococcal meningitis’clinical, biological and evolutive features in children]. Arch Pediatr. 2008;15 Suppl 3:S126–32.

26. Krause JC, Ghandil P, Chrabieh M, Casanova JL, Picard C, Puel A, et al. Very late-onset group B Streptococcus meningitis, sepsis, and systemic shigellosis due to interleukin-1 receptor-associated kinase-4 deficiency. Clin Infect Dis. 2009;49(9):1393–6.

27. Hosoda A, Gatayama R, Moriyama S, Ishii N, Yamada K, Matsuzaki Y, et al. The first case of recurrent ultra late onset group B streptococcal sepsis in a 3-year-old child. IDCases. 2017;7:16–8.

28. Takahara T, Matsubara K, Maihara T, Yamagami Y, Chang B. Ultra-late-onset Meningitis Caused by Serotype IX Group B Streptococcus. Pediatr Infect Dis J. 34. United States 2015. p. 801.

